# Weighting of risk factors for low birth weight: A linked routine data cohort study in Wales, UK

**DOI:** 10.1101/2022.03.23.22272809

**Authors:** Amrita Bandyopadhyay, Hope Jones, Michael Parker, Emily Marchant, Julie Evans, Charlotte Todd, Muhammad A. Rahman, James Healy, Tint Lwin Win, Ben Rowe, Simon Moore, Angela Jones, Sinead Brophy

**Affiliations:** National Centre for Population Health and Wellbeing Research, Swansea University Medical School, Wales, SA2 8PP, UK; Public Health Wales, Keir Hardie University Health Park, CF48 1BZ, Wales. UK; Cardiff School of Technologies, Cardiff Metropolitan University, Llandaff Campus, CF5 2YB, Wales, UK; Office for National Statistics, Government Buildings, Cardiff Road, Duffryn, Newport, NP10 8XG, UK; National Police Chiefs’ Council Lead for Mental Health and Age, 10 Victoria Street, London, SW1H 0NN, UK; School of Dentistry, Cardiff University, Cardiff, Wales, CF14 4XY, UK; Security, Crime, Intelligence Institute, Cardiff University, SPARK, Maindy Road, Cardiff, Wales, CF10 3AE, UK

**Keywords:** Low birth weight, Maternal health, Pregnancy interval, Data linkage, Cohort study

## Abstract

**Background:** Globally 20 million children are born with a birth weight below 2,500 grams every year which is considered as a low birthweight (LBW) baby. This study investigates the contribution of modifiable risk factors to inform activities that reduce the rates of LBW in Wales.

**Method:** The cohort (N = 693,377) was comprised of children born between 1^st^ January 1998 and 31^st^ December 2018 in Wales. A multivariable logistic regression model and a predictive model using decision tree were used to investigate the associations between the risk factors and LBW.

**Results:** The study found that non-singleton children had the highest risk of LBW (adjusted odds ratio 21.74, 95% confidence interval (CI) 21.09 – 22.40), followed by pregnancy interval less than one year (2.92, 95% CI 2.70 – 3.15), maternal diabetes (2.03, 95% CI 1.81 – 2.28), maternal hospital admission for anaemia (1.26, 95% CI 1.16 – 1.36), depression (1.58, 95% CI 1.43 – 1.75), serious mental illness (1.46, 95% CI 1.04 – 2.05), anxiety (1.22, 95% CI 1.08 – 1.38) and use of anti-depressant medication during pregnancy (1.92, 95% CI 1.20 – 3.07). Additional maternal risk factors include smoking (1.80, 95% CI 1.76 – 1.84), alcohol-related hospital admission (1.60, 95% CI 1.30 – 1.97), substance misuse (1.35, 95% CI 1.29 – 1.41), living in areas of high deprivation, and evidence of domestic abuse.

**Conclusion:** This work suggests that to address LBW, measures need to focus on improving maternal health, addressing pre-term births, promoting awareness of sufficient pregnancy interval, ensuring adequate support and resources for mother’s mental health and wellbeing.

**What is already known on this topic:**

- Each year 6.9% of live births are identified as low birth weight (LBW) in the UK.
- LBW children are at risk of poor cognitive development, which is associated with developmental disabilities and poor academic achievement in later life.
- The progress to reduce the LBW prevalence in high income regions (including Europe) is not satisfactory to meet World Health Organisation (WHO) LBW target by 2025.

**What this study adds:**

- This work has built an e-cohort using data-linkage across multiple routinely collected administrative datasets to investigate the risk factors of LBW for the population of Wales.
- Non-singleton children had 22 times higher risk of LBW than singleton children.
- Findings suggested that the most important factors to address to reduce the risk of LBW are multiple births, underlying maternal physical (diabetes, anaemia) and mental health, maternal smoking, and substance use (alcohol/drugs), adequate pregnancy interval, higher deprivation, and domestic abuse during pregnancy.

**How this study might affect research, practice, or policy:**

- This finding suggests that to address the LBW, adequate support is needed to the mother and their family when they are planning for and during pregnancy.
- Multiagency (doctors, midwives, security department) can liaise between them and can take necessary mitigative action to reduce the prevalence of LBW.

## Introduction

The World Health Organisation (WHO) defines low birth weight (LBW) as infants weighting less than 2,500 grams (5.5 pounds) irrespective of gestational age [1,2]. Latest figures show that each year around 53,000 live births (6.9%) are identified as LBW in the UK [3]. LBW is the result of intra-uterine growth restriction (less than 10th centile of weight for sex and gestational age), prematurity (gestational age less than 37 weeks), or a combination of both [4]. LBW can impair the baby’s cognitive development and lead to developmental disabilities and poor academic achievement [5]. Furthermore, LBW significantly increases the risk of perinatal and neonatal mortality and longstanding morbidity in early and later life [6]. Whilst there has been a reduction in mortality amongst preterm infants in the last two decades, the incidence of preterm birth has increased in many developed countries [6–8]. The increase is also associated with preterm delivery of multiple pregnancies, with medically indicated preterm birth 10 times more likely in multiple pregnancies than singleton births [9]. To address the global burden of LBW, the Sixty Fifth World Health Assembly Resolution 65.6 endorsed a comprehensive implementation plan to achieve a 30% reduction in LBW by 2025 [1]. A study conducted on the birth data from 148 countries of 195 United Nations’ member states indicated that there had been a 2.9% reduction in the LBW prevalence in 2015, compared to 2000 worldwide. However, there has not been any change in the LBW prevalence in high income regions (including Europe) and the progress is slower than required to meet the WHO LBW target by 2025 [10].

Existing research has found factors linked with mothers, such as age, high deprivation, and low academic qualification, are associated with increased odds of LBW [11,12]. Modifiable risk factors for LBW include inter-pregnancy interval [13], maternal physical [14–17] and mental health [18,19], and environmental exposures during pregnancy [20]. Studies have also shown numerous health behaviours, particularly smoking during pregnancy [21,22], alcohol, in which there is a dose-response relationship with LBW [24], and/or illicit drug use [23] are modifiable risk factors. Indirect (negative maternal behaviours, inadequate nutrition or prenatal care, and increased stress) or direct (physical assault, sexual trauma) experience of intimate partner abuse during pregnancy can lead to adverse infant outcomes, including LBW [25,26].

It is important to gain an understanding of these risk factors, particularly modifiable risk factors, so that resources and interventions can be scheduled effectively. Moreover, the wide range of risk factors cannot be addressed in isolation. Most of the risk factors that are strongly independently associated with LBW are correlated. This study aimed to understand the contributions of risk factors to the burden of LBW for the population of Wales, using traditional statistical methods and supervised machine learning models.

## Method

### Participants and linkage

The linked data cohort (N = 693,377) was comprised of children born in Wales between 1^st^ January 1998 and 31^st^ December 2018. The study population was identified in the National Community Child Health database (NCCHD), which is a local Child Health System database held by the National Health Service (NHS). The participants were linked to the Wales-wide administrative register, the Wales Demographic Service Dataset (WDS). Linkage was undertaken using an anonymised encrypted linkage key, the Anonymised Linking Field (ALF), in the Secure Anonymised Information Linkage (SAIL) Databank [27]. WDS provided the anonymised residential linking fields (RALFs), which is an encrypted residential address and its corresponding lower super output area (LSOA, small geographic areas with a population of approximately 1,500) when the child was born. LSOA was linked with the Welsh Index of Multiple Deprivation (WIMD) 2014, which is a measure of relative deprivation. The participants flow diagram is displayed in Supplementary Figure 1.

### Explanatory variables

The maternal variables related to a childbirth were obtained from NCCHD and Maternal Indicator Database (MID). The variables for maternal physical and mental health during pregnancy were obtained from primary care Welsh Longitudinal General Practice (WLGP) and hospital admissions dataset known as Patient Episode database in Wales (PEDW). The record of physical assault linked with mothers during pregnancy was obtained from PEDW. The substance misuse database (SMD) provided the information on alcohol and other drug abuse by the mother during pregnancy. Area type (urban/rural) and local authority (LA) under which they lived during the pregnancy and their overall and physical environment quantified in the WIMD were included in this study. The derived maternal variables include multiple birth flag (to distinguish between singleton and non-singleton), pregnancy interval, harmonised maternal smoking, and maternal weight. The variables and their sources have been described in Supplementary Table 1.

The impact of domestic abuse was examined using a subset of the study population (participants from Rhondda Cynon Taff born between June 2016 and 2018) with linked Public Protection Notification (PPN) dataset [28].

### Outcome variable

In this study a binary variable was created using the birth weight variable obtained from NCCHD.

- LBW = birth weight <2,500
- Not LBW (nLBW) = birth weight ≥2,500

### Statistical Analysis

It is known that gestational age is highly correlated with LBW. However, as the gestational age is only obtained at the point of birth, making it a non-modifiable risk factor, this study has not considered it as a predictor variable. The models were stratified by the multiple birth as this is one of the main predictors of LBW. The missing records in birth weight variable were removed from the analysis. Since there was around 15% missing data in maternal weight variable, the variable was imputed by the simple random imputation method [29]. The missing data in the other explanatory variables (less than 10%) were recoded as ‘Unknown’. The birth record for stillbirth and pregnancy interval of less than 22 weeks (as that is the minimum duration for a considerable gestation period) were also not considered for the statistical analysis. Data preparation including data linkage and data cleaning for this analysis was done on SAIL DB2 SQL platform. All statistical analyses were performed in R version 4.0.3.

A multivariable logistic regression (MLR) model was developed to identify the most important risk factors associated with LBW. The MLR model was built on the overall study population (whole Wales dataset) to identify the associations between all the explanatory and outcome variables.

A supervised machined learning classifier -decision tree (DT) model was developed to build a risk profile for LBW and test its predictive performance. Classification tree – DT models were constructed using RPART (Recursive Partitioning And Regression Trees) packages in R [30,31]. The algorithm recursively partitions the data into multiple sub-spaces to obtain the homogeneous final sub-space of predictor variables. For DT, the whole Wales data except for Rhondda Cynon Taff was used to train the model and prediction performance was evaluated on a test dataset which consisted of a sample of participants from the LA of Rhondda Cynon Taff. This LA was chosen because it had one of the highest rates of LBW in Wales and is an area which would benefit most from an accurate prediction model.

A separate data linkage was undertaken with a subset of the study population which was linked to the mother’s domestic abuse record from PPN dataset (the latter was only available for Rhonda Cynon Taff). Another adjusted MLR model was developed on this linked data to investigate the risk association for LBW.

### Ethical approval

This study had been approved by the SAIL Databank independent Information Governance Review Panel (IGRP) (project number 0916 – WECC Phase 4).

## Results

The study population consisted of 693,377 children of those 54,214 were from Rhondda Cynon Taff and 639,163 were from other LAs. The children from Rhondda Cynon Taff, which was later used as a test set for DT were well representative of the Welsh population (see Table 1). In the overall study population, 51.26% were boys, 96.92% were singleton and 90.38% children were born to term (gestational age between 37 and 42 weeks). 49.85% children were born as the first child in the family. Mothers of 0.48% children were admitted to hospital for diabetes and 0.09% had a GP visit for diabetes, 1.27% had depression, 1.52% with anxiety and 0.02% were on antidepressant medication during pregnancy. There were 1.26% and 21.51% children whose mothers had alcohol-related substance misuse and smoking record during pregnancy respectively. The average maternal age at birth of child and maternal weight was 28 years and 70.82 kg (after imputation) respectively and 63.68% of them were living in densely populated urban areas. Overall, 7.1% (8.26% in test set and 7% in other LAs) of children were born as LBW.

**Table 1:**
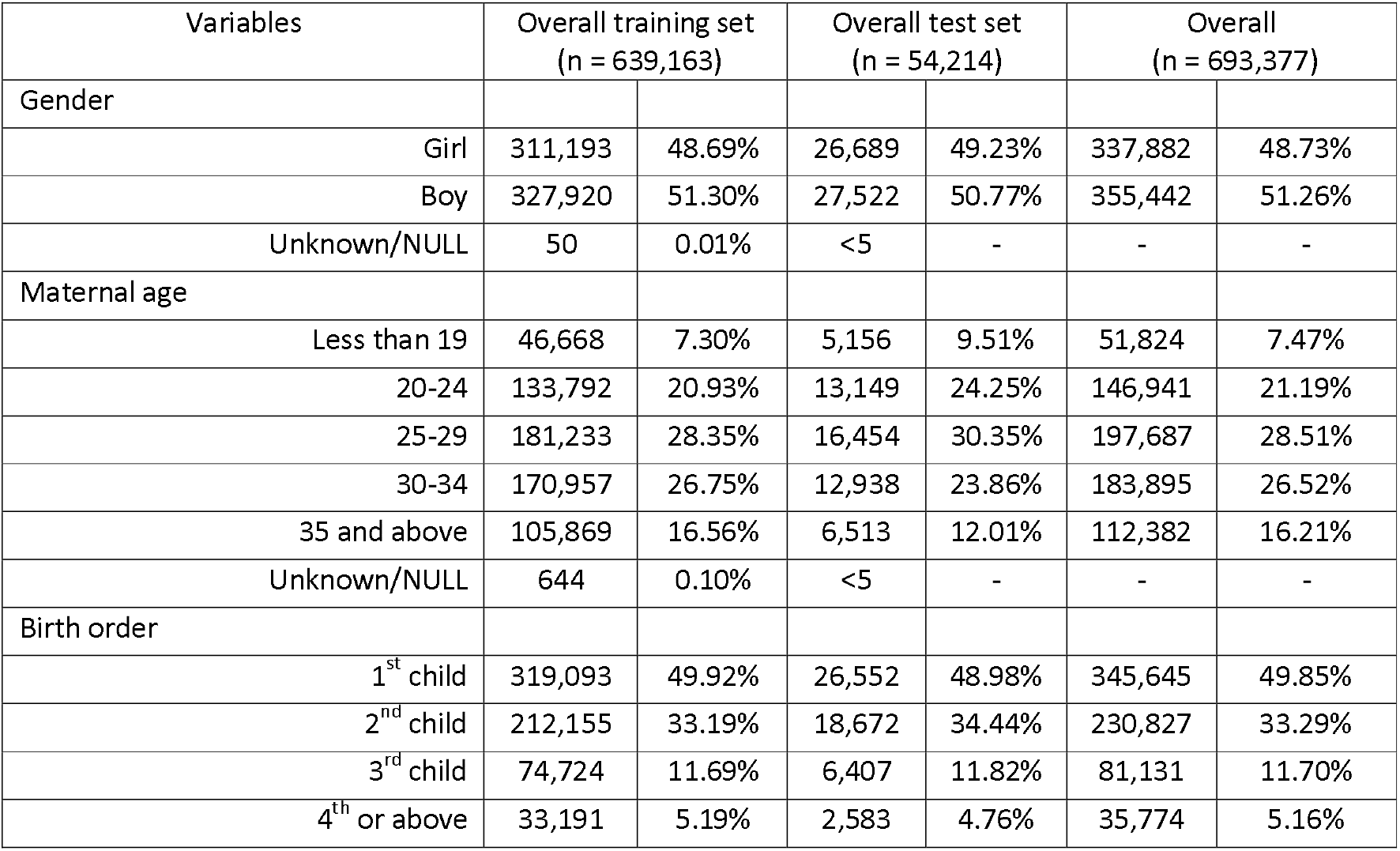
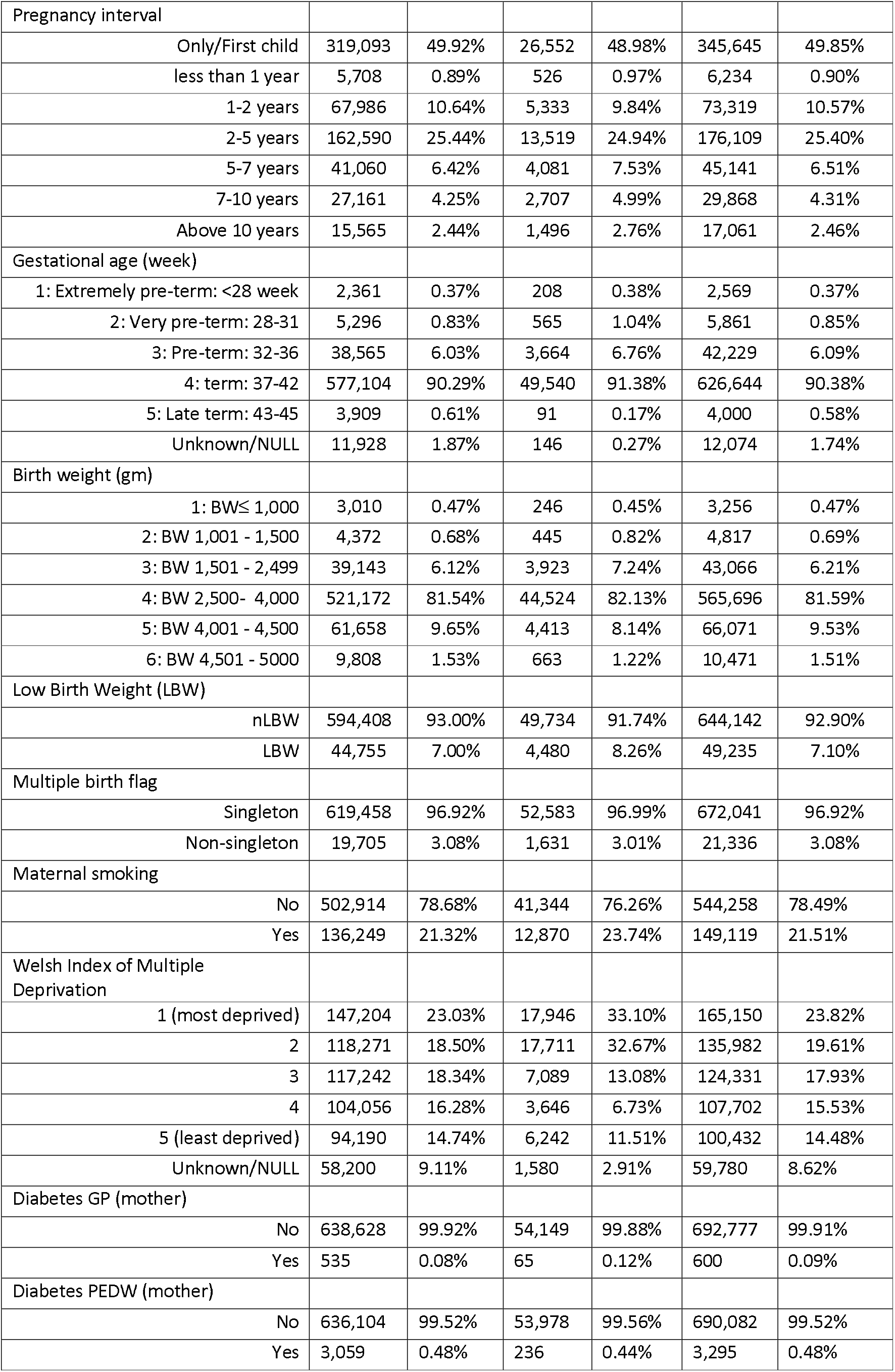
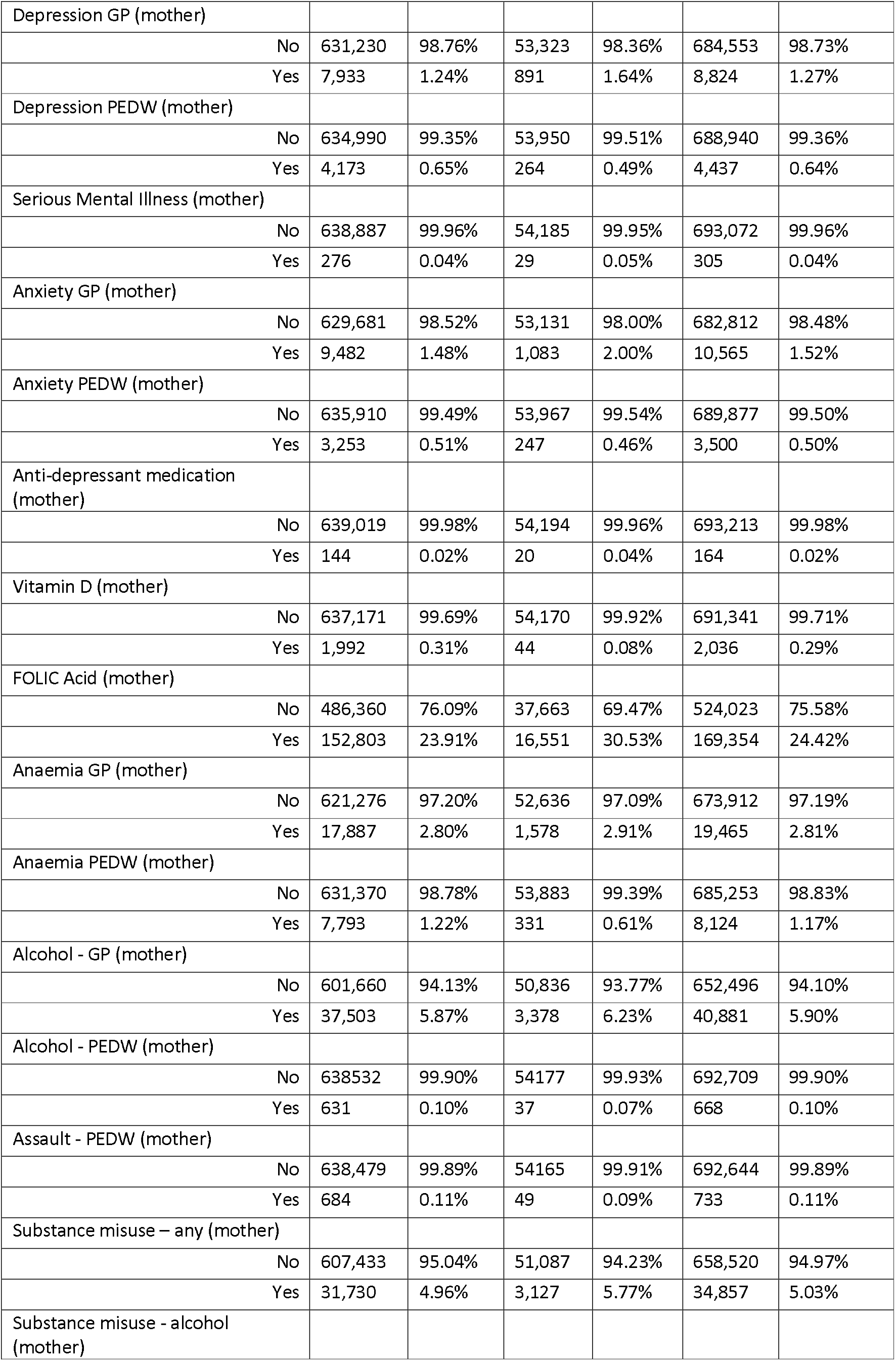
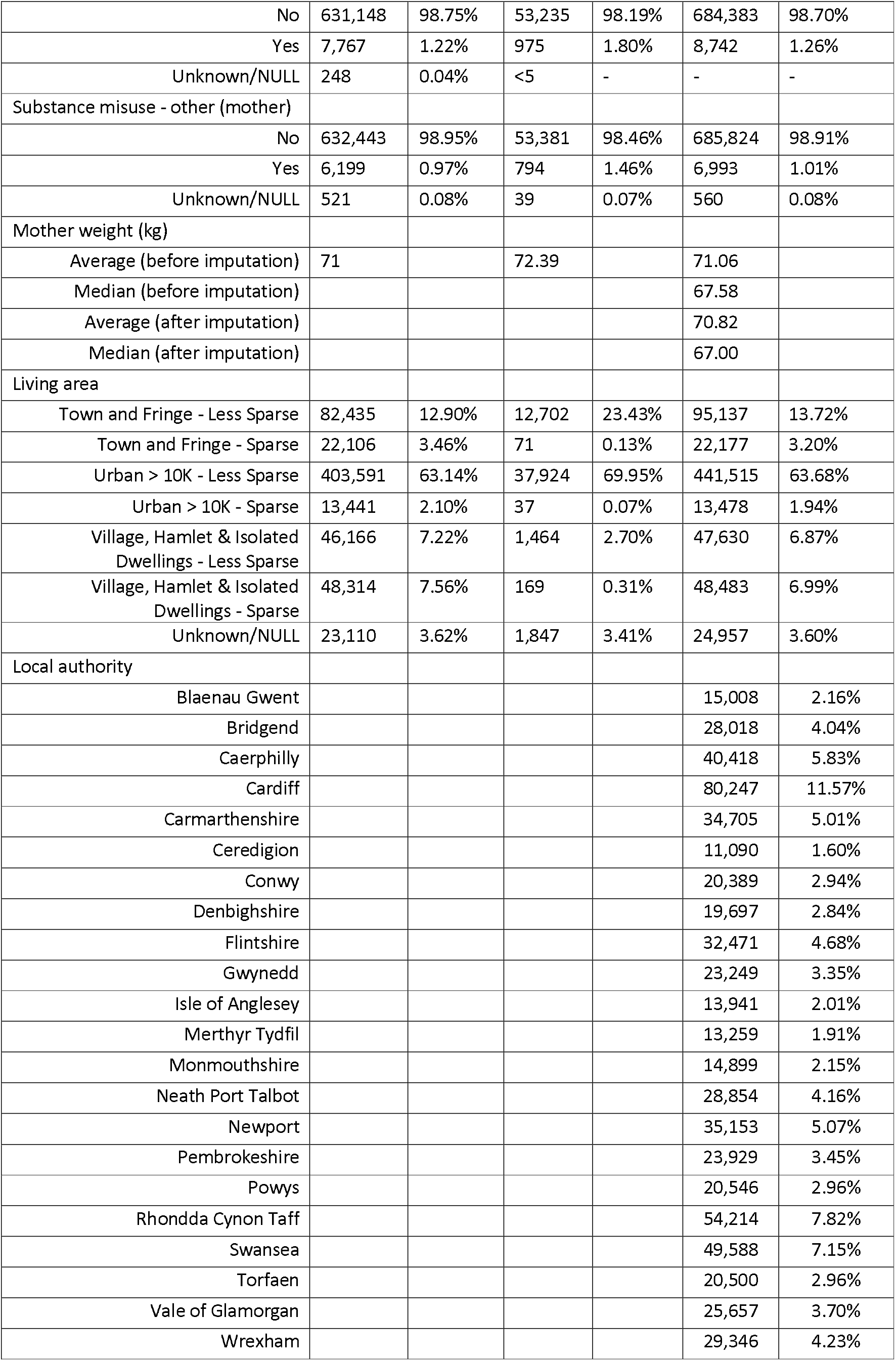
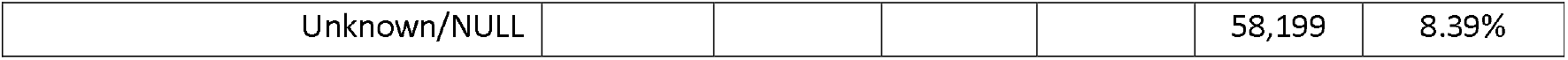
Characteristics of the study population.

### Factors associated with LBW: MLR results

Non-singleton children were at almost 22 times higher risk of LBW than singleton children (adjusted odds ratio (aOR) – 21.74 (95% confidence interval (CI) 21.09 – 22.40)). Mothers with diabetes-related GP visits (2.03 (1.81 – 2.28)) and hospital admission records of anaemia (1.26 (1.16 – 1.36)) during pregnancy were at very high risk of having LBW children. Poor mental health during pregnancy such as severe depression (1.58 (1.43– 1.75)), serious mental illness (1.46 (1.04 – 2.05)), severe anxiety (1.22 (1.08 – 1.38)) and antidepressant medications (1.92 (1.20– 3.07)) were risk factors for LBW. The other highly significant modifiable risk factors linked with pregnant mothers include maternal smoking (1.80 (1.76 – 1.84)), alcohol related hospital admissions (1.60 (1.30 – 1.97)) and any substance misuse (alcohol/other drugs) (1.35 (1.29 – 1.41)) during pregnancy. Higher maternal age was also associated with the risk of LBW. Though maternal age less than 19 was significantly associated with the risk of LBW in the univariable model, after adjusting all the other explanatory variables, this did not remain as a risk factor of LBW. The first child born was at higher risk of LBW than subsequent births, The odds of LBW for the 2^nd^ child was 0.59 (0.57 – 0.60) compared to the first child. Mothers living in the least deprived and rural areas during pregnancy were at lower risk of having LBW children than others living in more deprived and urban areas. The statistically significant risk factors with their aOR and CI have been described in Table 2 visualised in Supplementary Figure 2.

**Table 2:**
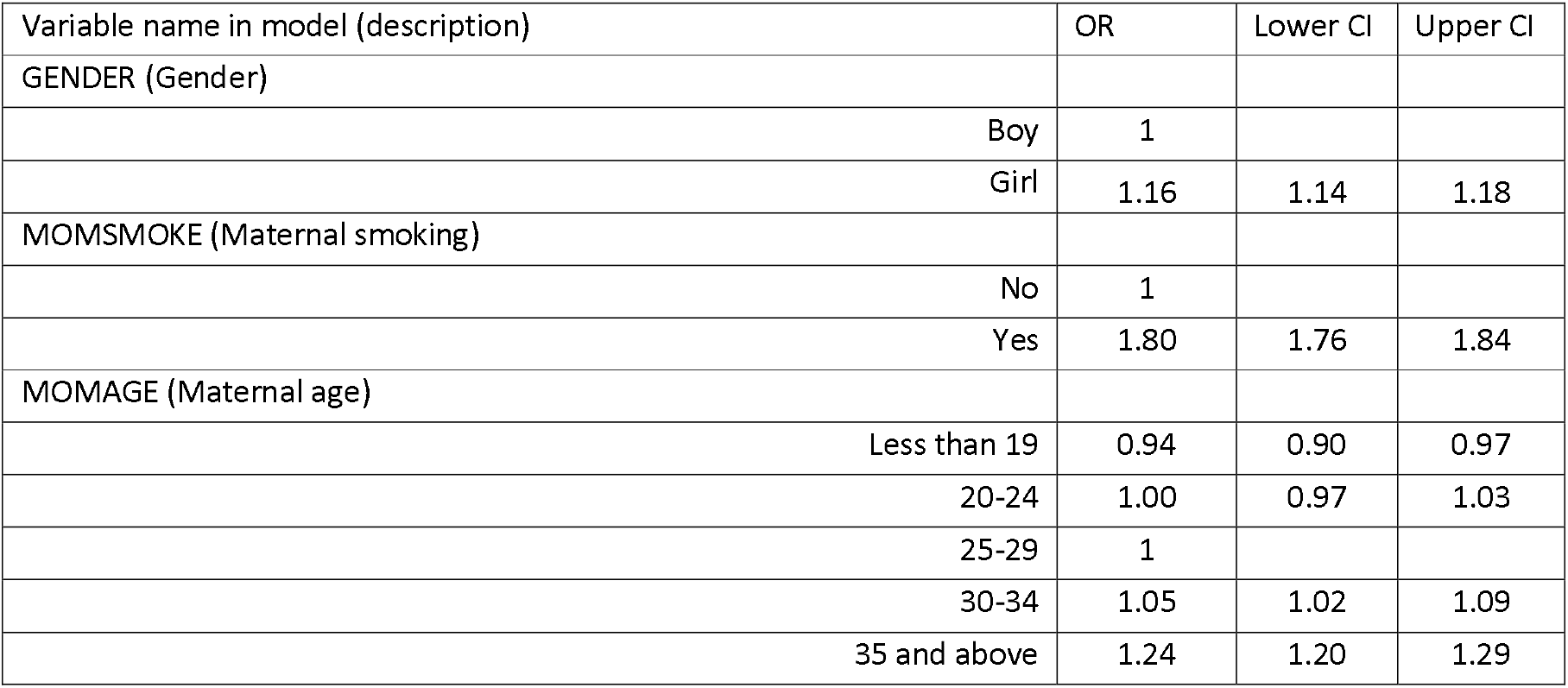
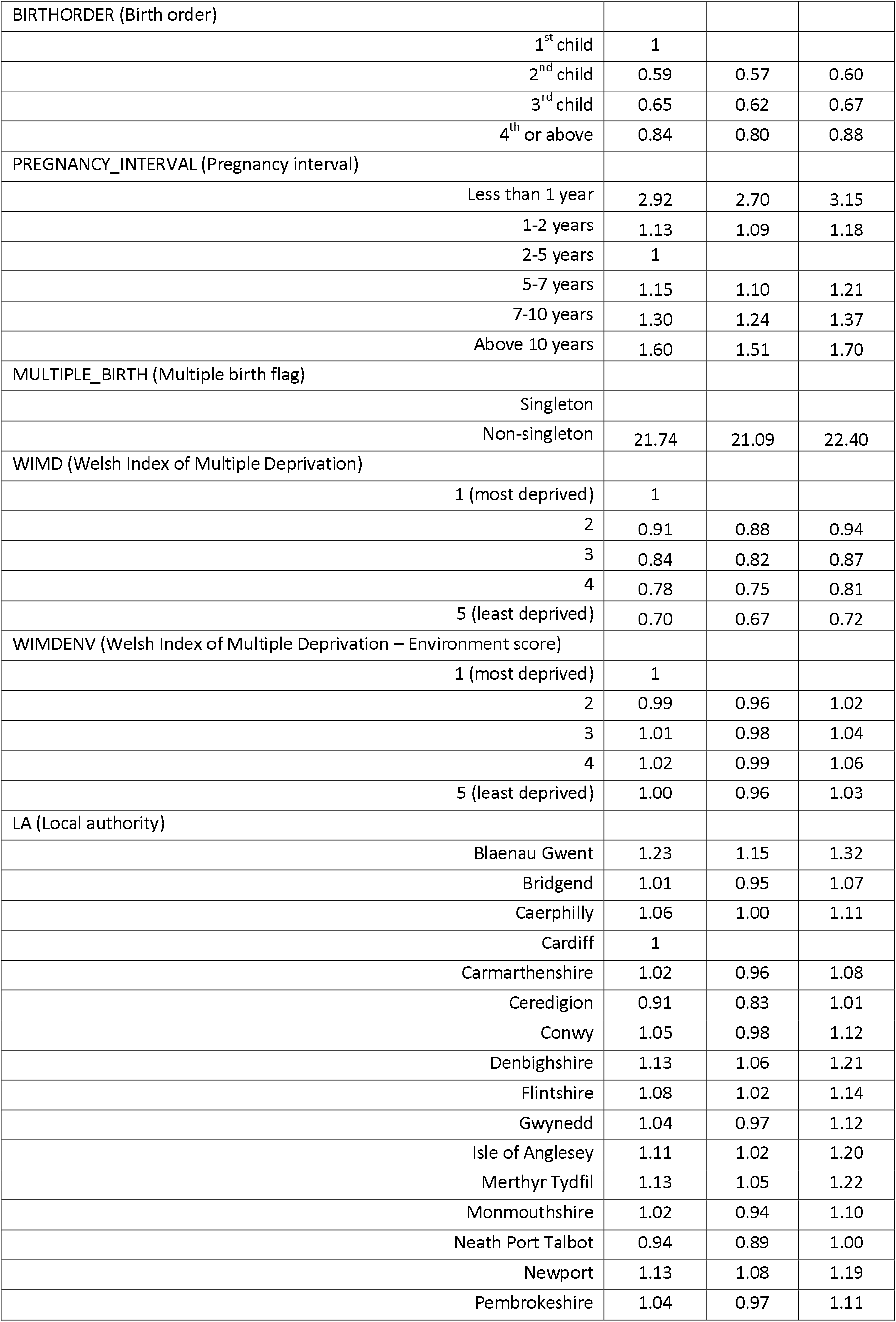
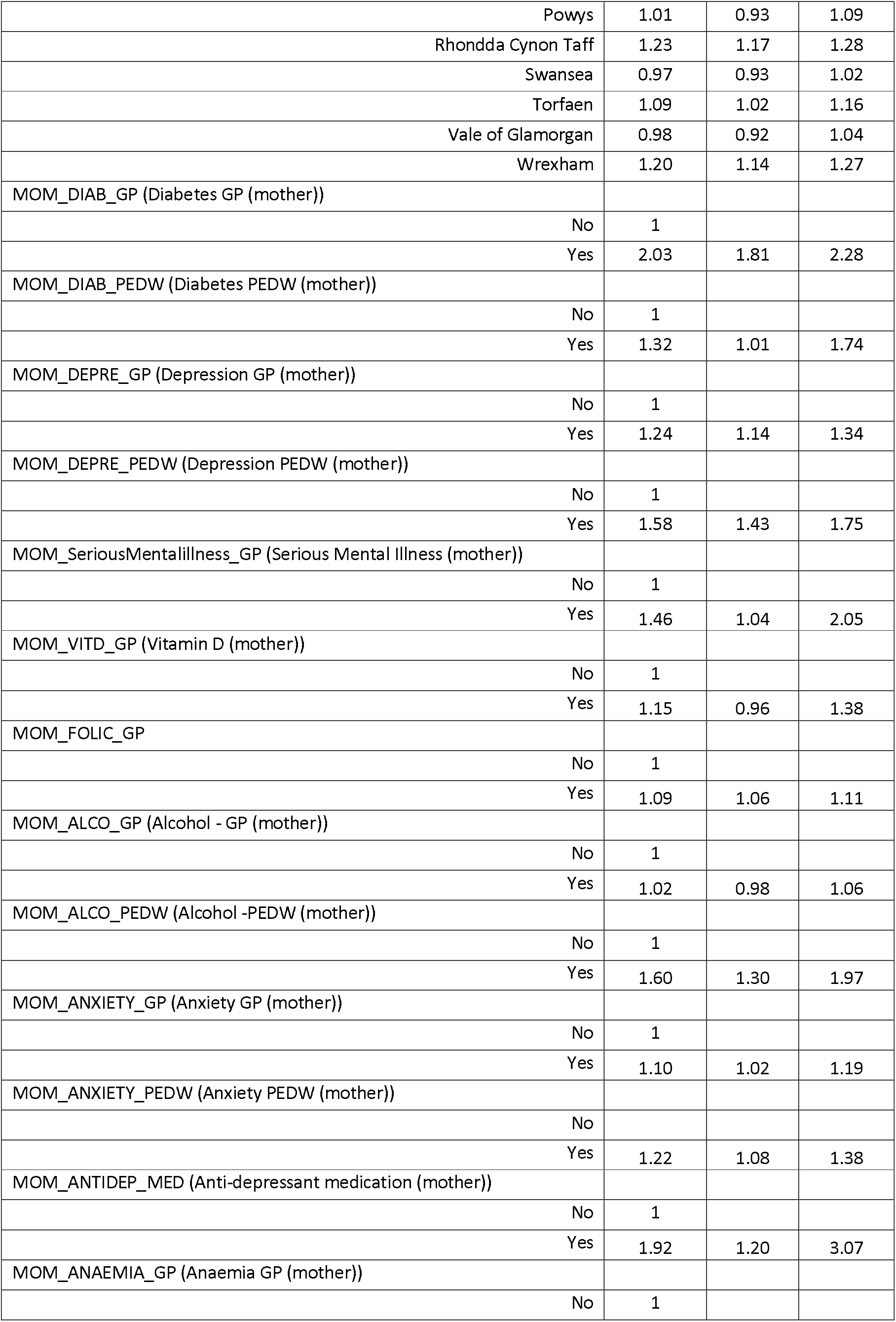
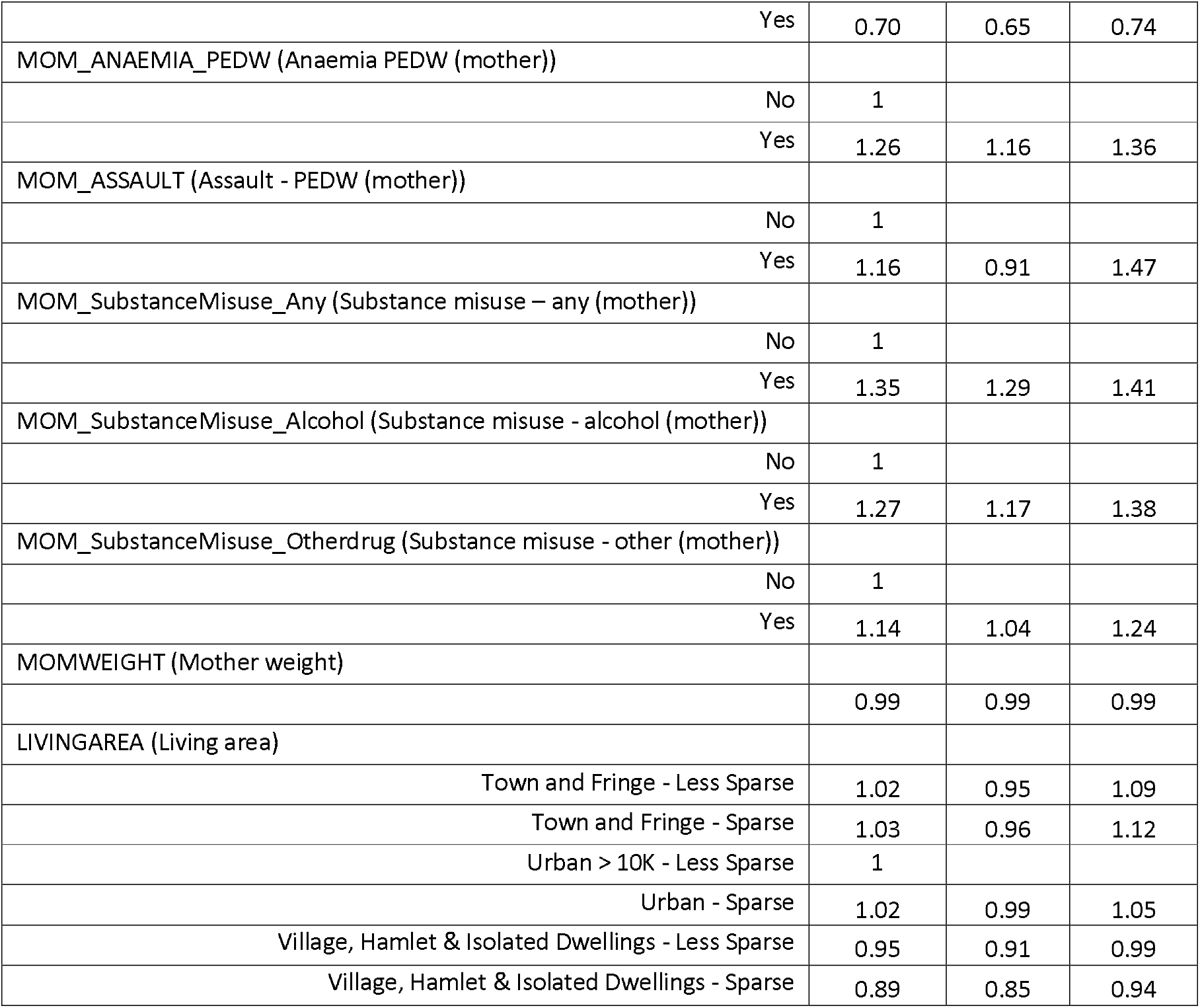
Multivariable logistic regression model to identify the risk factors of LBW among the overall study population.

### Finding from the linked PPN data model

Linkage of the cohort with PPN gave a dataset of 5,854 mothers of those who had a PPN call during pregnancy 18% had a LBW child whereas those who did not have PPN call 8.7% had a child with LBW (see Table 3). Mothers with a PPN call during pregnancy had almost 2 times higher risk of having a LBW baby (1.98 (1.39 – 2.81)) than mothers without PPN call after adjusting for confounding factors (see Supplementary Figure 5).

**Table 3:**
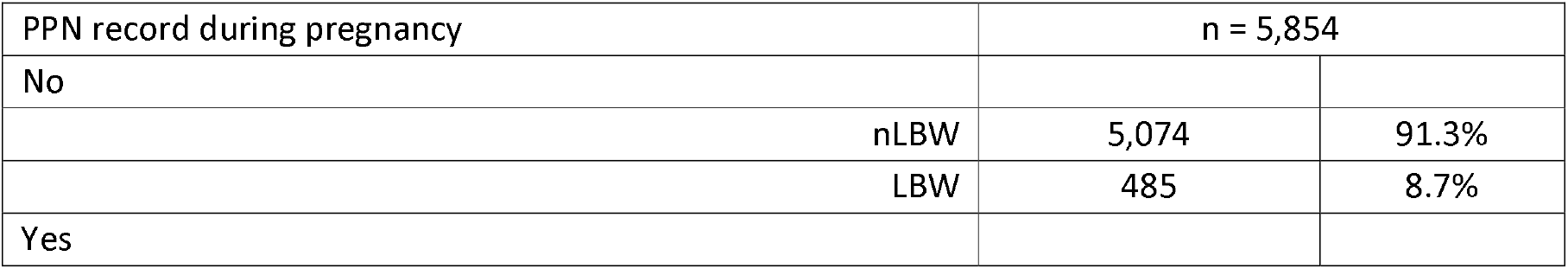
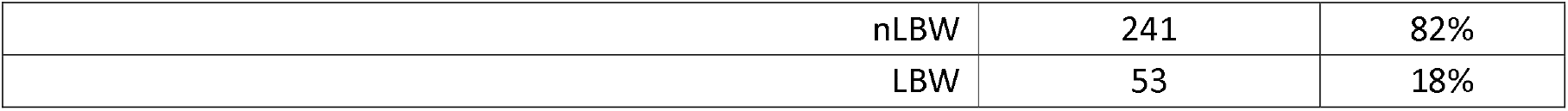
Distribution of LBW and nLBW children for the subset who were linked with mother’s PPN record during pregnancy.

### Predictive DT model

Since LBW were disproportionately more prevalent in non-singleton children (5.61% singleton vs 53.91% of the non-singleton children were LBW), two separate predictive models using DTs were developed.

### Singleton children

There were 619,458 observations in the training model. The most important risk factors selected by the DT algorithm to develop the final tree were maternal smoking, maternal weight, pregnancy interval, birth order, maternal substance misuse record (any), maternal age, deprivation - WIMD score, maternal substance misuse record (other drug) and maternal substance misuse record (alcohol). Supplementary Figure 3 depicts the final tree with the branches including final 33 terminal nodes. For example, the model would predict a LBW baby if a) maternal smoking is positive (e.g., mum smokes during pregnancy) and b) maternal weight less than 60 kg. The number of women in this category who had a LBW child is 73% (see terminal node 4 in Supplementary Figure 3) and risk profile was found in 7% of the training model population (e.g., 7% of pregnant women were smokers who weighed less than 60 kg during pregnancy).

The test data was built on the 52,583 singleton children, which is 7.82% of the total singleton children in this study. The model performance is explained in a confusion matrix with 60.54% accuracy, 60.41% sensitivity, 60.55% specificity, 9.68% positive predictive values and 95.63% negative predictive value (see Table 4 and 5).

**Table 4:** Confusion matrix/two by two table of the DT (singleton and non-singleton) models.

| Prediction | Reference<br>(Singleton)<br>n = 52,583 |  | Reference<br>(Non-singleton)<br>n = 1,631 |  |
| --- | --- | --- | --- | --- |
|  | LBW | nLBW | LBW | nLBW |
| LBW | 2,077 (TP) | 19,389 (FP) | 716 (TP) | 347 (FP) |
| nLBW | 1,361 (FN) | 29,756 (TN) | 326 (FN) | 242 (TN) |

**Table 5:**
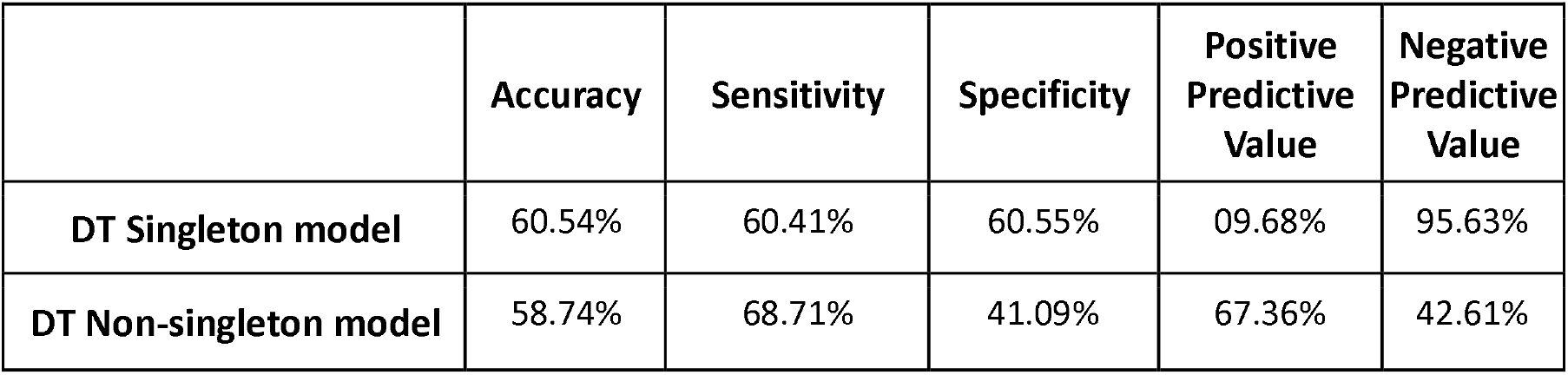
Prediction model performance (n=52,583 Singleton, n = 1,631 Non-singleton from test set)

|  | Accuracy | Sensitivity | Specificity | Positive Predictive Value | Negative Predictive Value |
| --- | --- | --- | --- | --- | --- |
| **DT Singleton model** | 60.54% | 60.41% | 60.55% | 09.68% | 95.63% |
| **DT Non-singleton model** | 58.74% | 68.71% | 41.09% | 67.36% | 42.61% |

### Non-singleton children

There were 19,705 children in the non-singleton training subset. The variables selected to generate the tree by the DT algorithm in the importance order were pregnancy interval, birth order, maternal weight, maternal age, gender, deprivation - WIMD score, maternal smoking, living area, deprivation - WIMD (environment) score and maternal substance misuse record (any). Figure 4 depicts the final tree with the branches including final 29 terminal nodes. For example, the model would predict a LBW baby if a) this is the first child or pregnancy interval is either above 10 years or less than 1 year and b) maternal weight less than 60 kg (terminal node 4).

The test set was built on the 1,631 non-singleton children, which is 7.64% of the total non-singleton children in this study. The model performance was measured as 58.74% accuracy, 68.71% sensitivity, 41.09% specificity, 67.36% positive predictive values and 42.61% negative predictive value (see Table 4 and 5).

## Discussion

7.1% of the overall study population in Wales was LBW between 1998 and 2018. Global trend of LBW is around 7.0% in both 2000 and 2015 for the developed regions (Europe, North America, Australia), which is consistent with our finding [2]. Findings from the Office for National Statistics (ONS) state a combined English and Welsh rate of LBW of 7.0% in 2016, unchanged from 2011 [32]. Our findings show that LBW is strongly associated with non-singleton pregnancy, and maternal health which includes a short pregnancy interval, non-optimal maternal body weight (e.g., low, or high weight), maternal smoking, diabetes, anaemia, mental illness and living in a deprived urban area and exposed to domestic abuse during pregnancy.

The findings of short and long pregnancy intervals being associated with increased odds of LBW has been reported previously [13]. However, Regan et al. highlighted many of the studies examining long inter-pregnancy interval are more prone to measurement error, with miscarriages and abortions in this time period difficult to capture and suggest caution should be taken interpreting these findings [33]. In terms of putting this evidence in context, when considering advice over pregnancy intervals, it will be important to consider all the available evidence including the impact of pregnancy interval on preterm birth and maternal outcomes [34]. Modifiable risk factors found to be important in this study included smoking during pregnancy. A number of reviews have been carried out in the field of interventions to reduce smoking in pregnancy which suggest that psychosocial interventions (counselling, feedback and incentives) appear to be effective at supporting women to stop smoking in pregnancy and can reduce the proportion of babies born LBW [35]. However, they argue that the context of the intervention needs to be given consideration and that whilst evidence exists for potentially effective interventions which could be piloted through delivery of programmes locally, efforts should also be directed at population wide strategies to reduce smoking uptake in young women. This may be especially important given the clear difficulties those pregnant experience in giving up smoking [35]. With regards to our finding of maternal mental health increasing the risk of LBW, both severe depression and anxiety were associated with an increased odds of LBW in our study [36].

The singleton DT model correctly predicted 60.41% of all the true positive cases. However, the positive predictive value of 9.68% indicates that the model assigned a false positive ‘LBW’ classification for 89.32% cases. This model only includes singleton children and since non-singleton pregnancies are highly associated with LBW, removing this variable from the model has lessened its predictive capability. This is evidenced by the significantly improved positive predictive value (67.36%) for the non-singleton model (table 5). Previous machine learning models appear to show better prediction as they included non-singleton, gestational age (which is in terms of temporal association highly associated with LBW but occurs at the same time as the LBW can be measured) and preeclampsia in third trimester.

The strength of this study lies in using national datasets of all births in Wales across a large time. However, as this was a linked routine data study some of the lifestyle factors cannot be captured (diet, physical activity, stress, emotional state) which can be important in determining LBW [37,38]. This work can only identify the more severe cases which are recorded in the health care system, and undiagnosed cases that did not result in the system will be missed. The two different models (MLR and DT) used in this study found very similar findings suggesting that factors which are common and so are predictive (using DT methods) such as maternal smoking status and low weight mother could be targeted to address population risk. Factors which have a strong association with LBW (using regression analysis) such as a mother with diabetes or mother on antidepressants, can be addressed to reduce individual risk for that mother/child.

## Conclusion

This study suggests that the most important factors to reduce the risk of LBW are to address multiple birth (e.g. in assisted reproduction practices), addressing factors associated with preterm births (previous history of preterm birth), addressing maternal health such as reducing smoking, investment in maternal mental health, addressing substance use (alcohol/drugs), treating underlying health conditions (diabetes/anaemia), and promoting planning of pregnancy to give an adequate pregnancy interval and healthy weight of mother especially for those in deprived urban areas.

## Data Availability

The linked data have been archived in Secure Anonymised Information Linkage (SAIL) Databank. They are available upon reasonable request to the SAIL databank.
https://duckduckgo.com/?t=ffab&q=SAIL++databank&ia=web

## Funding

This work was funded by Public Health Wales (PHW), grant number (105186). This work was supported by Nation Institute for Health Research (NIHR), grant number (NIHR133680). This research has been carried out as part of the ADR Wales programme of work. The ADR Wales programme of work is aligned to the priority themes as identified in the Welsh Government’s national strategy: Prosperity for All. ADR Wales brings together data science experts at Swansea University Medical School, staff from the Wales Institute of Social and Economic Research, Data and Methods (WISERD) at Cardiff University and specialist teams within the Welsh Government to develop new evidence which supports Prosperity for All by using the SAIL Databank at Swansea University, to link and analyse anonymised data. ADR Wales is part of the Economic and Social Research Council (part of UK Research and Innovation) funded ADR UK (grant ES/S007393/1).

This work was also supported by the National Centre for Population Health and Well-Being Research (NCPHWR) which is funded by Health and Care Research Wales. This work was supported by Health Data Research UK which receives its funding from HDR UK Ltd (NIWA1) funded by the UK Medical Research Council, Engineering and Physical Sciences Research Council, Economic and Social Research Council, Department of Health and Social Care (England), Chief Scientist Office of the Scottish Government Health and Social Care Directorates, Health and Social Care Research and Development Division (Welsh Government), Public Health Agency (Northern Ireland), British Heart Foundation (BHF) and the Welcome Trust.

This work uses data provided by patients and collected by the NHS as part of their care and support. This study used anonymised data held in the Secure Anonymised Information Linkage (SAIL) Databank. We would like to acknowledge all the data providers who enable SAIL to make anonymised data available for research. We acknowledge the support provided by South Wales Police.

## Contributorship statement

All authors contributed to the study conception and design. The police PPN and MID data accusation was supported by Ben Rowe and Julie Evans respectively. The core dataset was prepared by Muhammad A. Rahman. Further data preparation and the full analysis was done by Amrita Bandyopadhyay. James Healy, and Michael Parker contributed to the analysis. The first draft of the manuscript was written by Amrita Bandyopadhyay and all authors commented on previous versions of the manuscript. All authors read and approved the final manuscript.

### Conceptualization

Sinead Brophy, Angela Jones and Julie Evans and Amrita Bandyopadhyay; Methodology: Amrita Bandyopadhyay and Sinead Brophy; Formal analysis and investigation: Amrita Bandyopadhyay Writing - original draft preparation: Amrita Bandyopadhyay; Writing - review and editing: Charlotte Todd, Michael Parker, Julie Evans, Emily Marchant, Hope Jones, Muhammad A. Rahman, James Healy, Tint Lwin Win, Ben Rowe, Simon Moore, Angela Jones, and Sinead Brophy, Supervision: Sinead Brophy.

## Refence

1 WHO | Global Nutrition Targets 2025: Low birth weight policy brief. WHO. http://www.who.int/nutrition/publications/globaltargets2025_policybrief_lbw/en/ (accessed 19 Nov 2020).

2 UNICEF-WHO Low birthweight estimates: Levels and trends 2000–2015. https://www.unicef.org/reports/UNICEF-WHO-low-birthweight-estimates-2019 (accessed 3 Mar 2022).

3 Johnson CD, Jones S, Paranjothy S. Reducing low birth weight: prioritizing action to address modifiable risk factors. J Public Health Oxf Engl 2017;39:122–31. doi:10.1093/pubmed/fdv212

4 Mohammed SG. Low birth weight in Omdurman maternity hospital. Int J Sci Res Publ 2014;4:1– 13.

5 Breslau N, Paneth NS, Lucia VC. The Lingering Academic Deficits of Low Birth Weight Children. Pediatrics 2004;114:1035–40. doi:10.1542/peds.2004-0069

6 Ohlsson A, Shah P. Determinants and prevention of low birth weight: a synopsis of the evidence. Institute of Health Economics 2008.

7 Heaman MI, Sprague AE, Stewart PJ. Reducing the Preterm Birth Rate: A Population Health Strategy. J Obstet Gynecol Neonatal Nurs 2001;30:20–9. doi:https://doi.org/10.1111/j.1552-6909.2001.tb01518.x

8 Yuan W, Duffner AM, Chen L, et al. Analysis of preterm deliveries below 35 weeks’ gestation in a tertiary referral hospital in the UK. A case-control survey. BMC Res Notes 2010;3:119. doi:10.1186/1756-0500-3-119

9 Blencowe H, Cousens S, Chou D, et al. Born Too Soon: The global epidemiology of 15 million preterm births. Reprod Health 2013;10:S2. doi:10.1186/1742-4755-10-S1-S2

10 Blencowe H, Krasevec J, Onis M de, et al. National, regional, and worldwide estimates of low birthweight in 2015, with trends from 2000: a systematic analysis. Lancet Glob Health 2019;7:e849–60. doi:10.1016/S2214-109X(18)30565-5

11 Shi L, Macinko J, Starfield B, et al. Primary care, infant mortality, and low birth weight in the states of the USA. J Epidemiol Community Health 2004;58:374–80. doi:10.1136/jech.2003.013078

12 Silvestrin S, Silva CH da, Hirakata VN, et al. Maternal education level and low birth weight: a meta-analysis. J Pediatr (Rio J) 2013;89:339–45. doi:10.1016/j.jped.2013.01.003

13 Conde-Agudelo A, Rosas-Bermúdez A, Kafury-Goeta AC. Birth Spacing and Risk of Adverse Perinatal Outcomes: A Meta-analysis. JAMA 2006;295:1809–23. doi:10.1001/jama.295.15.1809

14 Yu Z, Han S, Zhu J, et al. Pre-Pregnancy Body Mass Index in Relation to Infant Birth Weight and Offspring Overweight/Obesity: A Systematic Review and Meta-Analysis. PLOS ONE 2013;8:e61627. doi:10.1371/journal.pone.0061627

15 Daalderop LA, Wieland BV, Tomsin K, et al. Periodontal Disease and Pregnancy Outcomes: Overview of Systematic Reviews. JDR Clin Transl Res 2018;3:10–27. doi:10.1177/2380084417731097

16 Flynn CA, Helwig AL, Meurer LN. Bacterial vaginosis in pregnancy and the risk of prematurity. J Fam Pract 1999;48:885–92.

17 Figueiredo ACMG, Gomes-Filho IS, Silva RB, et al. Maternal Anemia and Low Birth Weight: A Systematic Review and Meta-Analysis. Nutrients 2018;10. doi:10.3390/nu10050601

18 Dadi AF, Miller ER, Bisetegn TA, et al. Global burden of antenatal depression and its association with adverse birth outcomes: an umbrella review. BMC Public Health 2020;20:173. doi:10.1186/s12889-020-8293-9

19 Lima SAM, Dib RPE, Rodrigues MRK, et al. Is the risk of low birth weight or preterm labor greater when maternal stress is experienced during pregnancy? A systematic review and meta-analysis of cohort studies. PLOS ONE 2018;13:e0200594. doi:10.1371/journal.pone.0200594

20 Fleischer Nancy L., Merialdi Mario, van Donkelaar Aaron, et al. Outdoor Air Pollution, Preterm Birth, and Low Birth Weight: Analysis of the World Health Organization Global Survey on Maternal and Perinatal Health. Environ Health Perspect 2014;122:425–30. doi:10.1289/ehp.1306837

21 Flower A, Shawe J, Stephenson J, et al. Pregnancy planning, smoking behaviour during pregnancy, and neonatal outcome: UK millennium cohort study. BMC Pregnancy Childbirth 2013;13:238. doi:10.1186/1471-2393-13-238

22 Jaddoe VWV, Troe E-JWM, Hofman A, et al. Active and passive maternal smoking during pregnancy and the risks of low birthweight and preterm birth: the Generation R Study. Paediatr Perinat Epidemiol 2008;22:162–71. doi:10.1111/j.1365-3016.2007.00916.x

23 dos Santos JF, de Melo Bastos Cavalcante C, Barbosa FT, et al. Maternal, fetal and neonatal consequences associated with the use of crack cocaine during the gestational period: a systematic review and meta-analysis. Arch Gynecol Obstet 2018;298:487–503. doi:10.1007/s00404-018-4833-2

24 Patra J, Bakker R, Irving H, et al. Dose–response relationship between alcohol consumption before and during pregnancy and the risks of low birthweight, preterm birth and small for gestational age (SGA)—a systematic review and meta-analyses. BJOG Int J Obstet Gynaecol 2011;118:1411–21. doi:10.1111/j.1471-0528.2011.03050.x

25 Hill A, Pallitto C, McCleary-Sills J, et al. A systematic review and meta-analysis of intimate partner violence during pregnancy and selected birth outcomes. Int J Gynaecol Obstet Off Organ Int Fed Gynaecol Obstet 2016;133:269–76. doi:10.1016/j.ijgo.2015.10.023

26 Donovan BM, Spracklen CN, Schweizer ML, et al. Intimate partner violence during pregnancy and the risk for adverse infant outcomes: a systematic review and meta-analysis. BJOG Int J Obstet Gynaecol 2016;123:1289–99. doi:https://doi.org/10.1111/1471-0528.13928

27 Lyons RA, Jones KH, John G, et al. The SAIL databank: linking multiple health and social care datasets. BMC Med Inform Decis Mak 2009;9:3. doi:10.1186/1472-6947-9-3

28 Constabulary © Her Majesty’s Inspectorate of, Fire. Police effectiveness 2015 (Vulnerability) – Dyfed-Powys Police. HMICFRS. https://www.justiceinspectorates.gov.uk/hmicfrs/publications/police-effectiveness-vulnerability-2015-dyfed-powys/ (accessed 3 Nov 2021).

29 Gelman A, Hill J. Data analysis using regression and multilevel/hierarchical models. Cambridge university press 2006.

30 Lewis RJ. An introduction to classification and regression tree (CART) analysis. In: Annual meeting of the society for academic emergency medicine in San Francisco, California. 2000.

31 Atkinson Beth. rpart function | R Documentation. https://www.rdocumentation.org/packages/rpart/versions/4.1-15/topics/rpart (accessed 14 Jan 2021).

32 ONS. Birth characteristics in England and Wales - Office for National Statistics. 2019.https://www.ons.gov.uk/peoplepopulationandcommunity/birthsdeathsandmarriages/livebirths/bulletins/birthcharacteristicsinenglandandwales/2017 (accessed 14 Jul 2021).

33 Regan AK, Ball SJ, Warren JL, et al. A Population-Based Matched-Sibling Analysis Estimating the Associations Between First Interpregnancy Interval and Birth Outcomes. Am J Epidemiol 2019;188:9–16. doi:10.1093/aje/kwy188

34 Hutcheon JA, Nelson HD, Stidd R, et al. Short interpregnancy intervals and adverse maternal outcomes in high-resource settings: An updated systematic review. Paediatr Perinat Epidemiol 2019;33:O48–59. doi:10.1111/ppe.12518

35 Chamberlain C, O’Mara-Eves A, Porter J, et al. Psychosocial interventions for supporting women to stop smoking in pregnancy. Cochrane Database Syst Rev Published Online First: 2017. doi:10.1002/14651858.CD001055.pub5

36 Howard LM, Khalifeh H. Perinatal mental health: a review of progress and challenges. World Psychiatry 2020;19:313–27. doi:10.1002/wps.20769

37 Ghavi A, Fadakar Sogheh K, Niknamy M, et al. Investigating the Relationship between Maternal Lifestyle during Pregnancy and Low-Birth-Weight of Term Neonates. Iran J Obstet Gynecol Infertil 2012;15:14–24.

38 Xi C, Luo M, Wang T, et al. Association between maternal lifestyle factors and low birth weight in preterm and term births: a case-control study. Reprod Health 2020;17:93. doi:10.1186/s12978-020-00932-9

