## Supplemental figures for "Weighting of risk factors for low birth weight: A linked routine data cohort study in Wales, UK"

**Supplementary Figure 1 Participants flow diagram.**

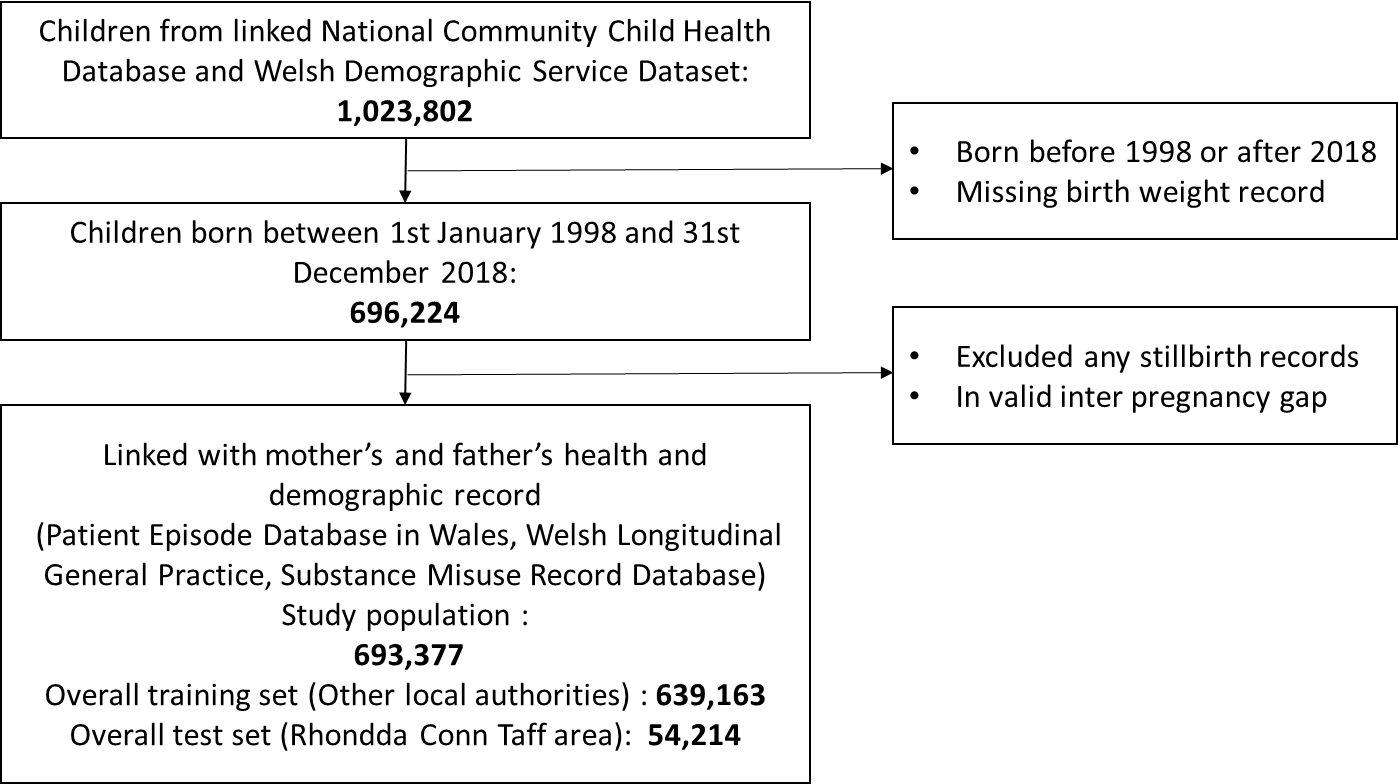

**Supplementary Figure 2: Significant factors associated with the risk LBW among the overall study population.**

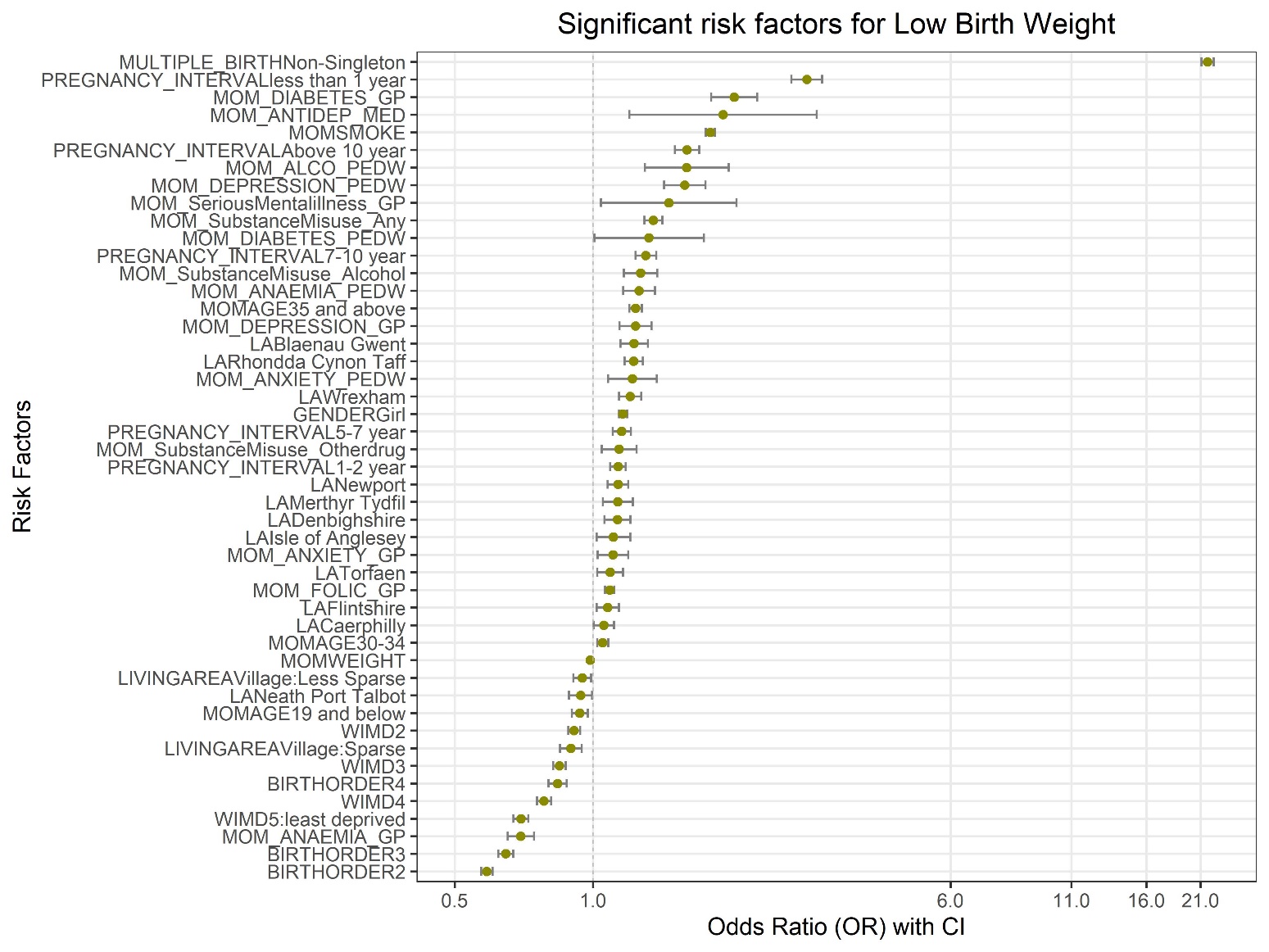

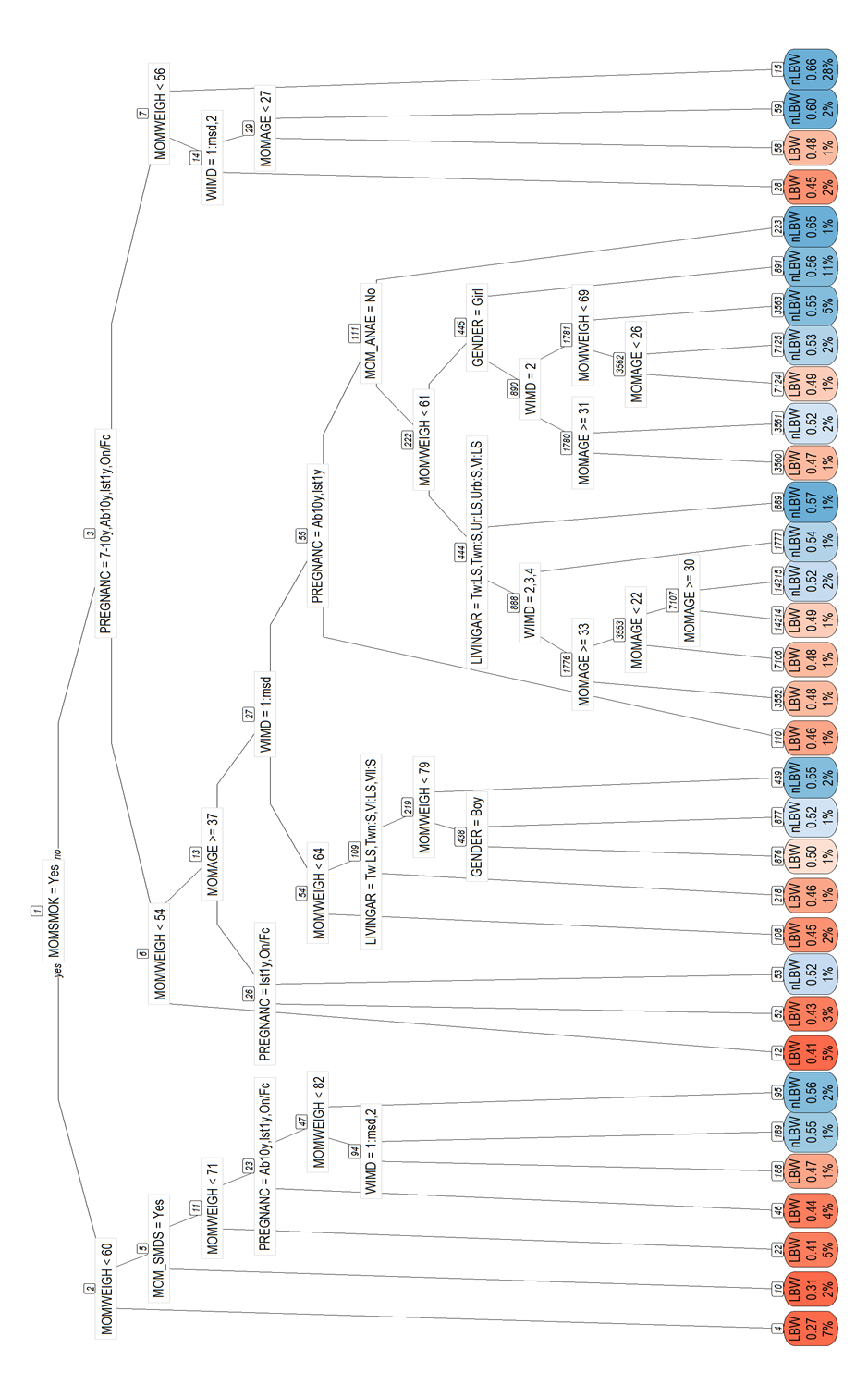

**Supplementary Figure 3: Decision tree for singleton children**

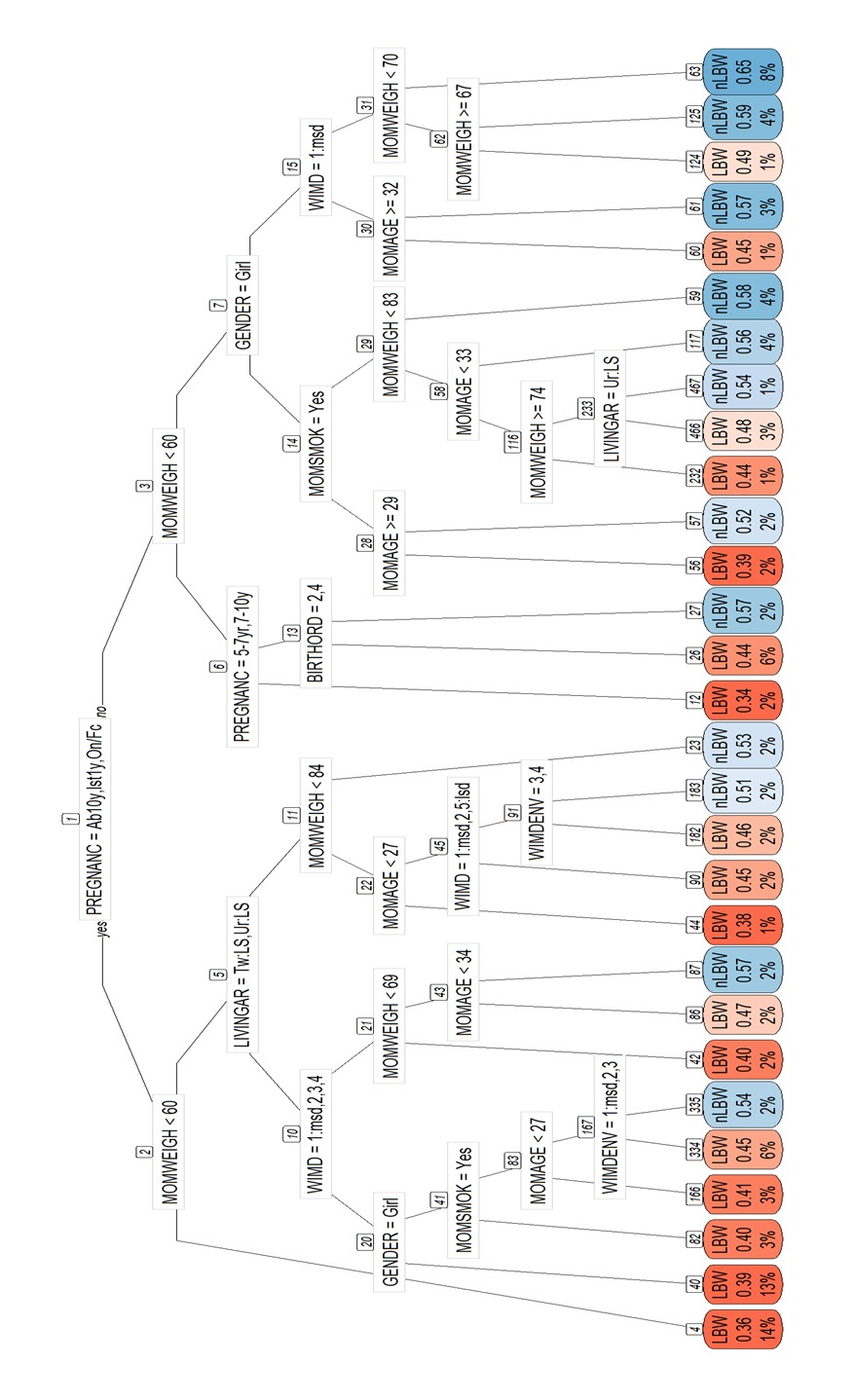

**Supplementary Figure 4: Decision tree for non-singleton children**

**Supplementary Figure 5: Significant risk factors associated with the risk LBW after linking with PPN record**

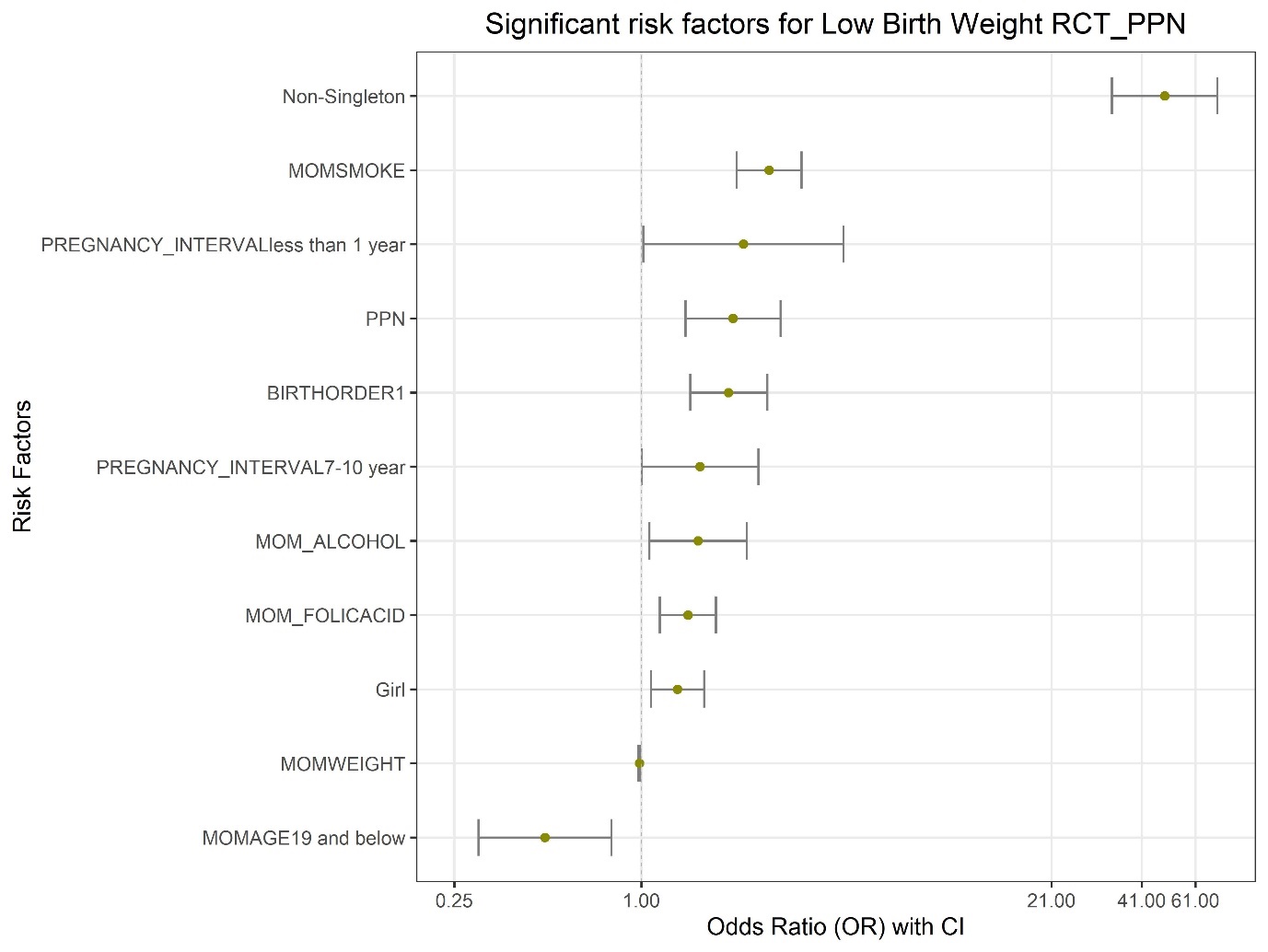
