## Supplemental tables for "Weighting of risk factors for low birth weight: A linked routine data cohort study in Wales, UK"

**Supplementary Table1: Variables and their source datasets**

| Variables | NCCHD | MID | WLGP | PEDW | WDS | SMD | Derived | Description if derived |
| --- | --- | --- | --- | --- | --- | --- | --- | --- |
| Gender | + |  |  |  |  |  |  |  |
| Maternal age | + |  |  |  |  |  |  |  |
| Gestational age | + |  |  |  |  |  |  |  |
| Birth weight | + |  |  |  |  |  |  |  |
| Birth order | + |  |  |  |  |  |  |  |
| Pregnancy interval |  |  |  |  |  |  | + | Pregnancy interval, in week format, was derived using the birth order, week of birth (the Monday of the week of date of birth), of the previous child and the current child, maternal identifier, and the multiple birth flag. |
| Multiple birth flag |  |  |  |  |  |  | + | Using the week of birth, encrypted maternal identifier and the birth order, a binary variable – ‘multiple birth flag’ was derived to distinguish between singleton and non-singleton birth. |
| Mother weight (kg) |  |  |  |  |  |  | + | The maternal weight during pregnancy was obtained from MID and WLGP. The final maternal weight variable was derived following cleaning and harmonising it with the source variables which includes removing and recoding missing, erroneous, and inconsistent records. |
| Maternal smoking | + | + | + |  |  |  |  | A cleaned and harmonised variable of maternal smoking during pregnancy was created based on the data obtained from three sources. |
| WIMD |  |  |  |  | + |  |  |  |
| Diabetes |  |  | + | + |  |  |  |  |
| Depression |  |  | + | + |  |  |  |  |
| Serious Mental Illness |  |  | + |  |  |  |  |  |
| Anxiety |  |  | + | + |  |  |  |  |
| Anti-depressant medication |  |  | + |  |  |  |  |  |
| Vitamin D |  |  | + |  |  |  |  |  |
| FOLIC Acid |  |  | + |  |  |  |  |  |
| Anaemia |  |  | + | + |  |  |  |  |
| Alcohol |  |  | + | + |  |  |  |  |
| Assault |  |  |  | + |  |  |  |  |
| Substance misuse |  |  |  |  |  | + |  |  |
| Living area |  |  |  |  | + |  |  |  |
| Local authority |  |  |  |  | + |  |  |  |

**Supplementary Table 2: Distribution of LBW and nLBW children based on their multiple birth flags**

|  | Overall training set  (n = 639,163) | | Overall test set  (n = 54,214) | |
| --- | --- | --- | --- | --- |
| Singleton |  |  |  |  |
| nLBW | 585,163 | 94.46% | 49,145 | 93.46% |
| LBW | 34,295 | 5.54% | 3,438 | 6.54% |
| Non-singleton |  |  |  |  |
| nLBW | 9,245 | 46.92% | 589 | 36.11% |
| LBW | 10,460 | 53.08% | 1,042 | 63.89% |
